# The implications of religious event and holidays on the transmission of SARS-CoV2 : the impact of behavioral changes on transmission

**DOI:** 10.1101/2020.10.17.20214056

**Authors:** Abdulkarim Abdulrahman, Muath AlMajthoob, Abdulla I AlAwadhi, Manaf M AlQahtani

## Abstract

**Introduction:** The risk factors for transmission of SARS-CoV2 have been widely studied and it was evident that a population’s behavior has a direct effect on the risk of transmission. Public health measures and regulation are largely kept to control and direct these behaviors. In this study, we describe the change in transmission in SARS-CoV2 in relation to demographics before and after two major religious events: “Eid Alfitr” and “Ashura”

**Methods:** This is a national observational study conducted in Bahrain in September 2020 to compare the number and demographics of all newly diagnosed cases before and after Eid Alfitr (religious holiday) and Ashura religious event. A 10 day period before the event was compared to a 10 day period after the event by ten days. Data on the number of tests, number of new cases, their demographics (age, gender, nationality) and presence of symptoms were collected and analyzed.

**Results:** There was significant increase in the number of COVID-19 cases after both Eid Alfitr (1997 more cases, with a 67% increase) and Ashura (4232 more cases with 2.19 times more cases). The majority of new cases after the religious events were found in local Bahrainis, from 472 cases to 2169 cases after Eid, and from 2201 to 6639 cases after Ashura. The rise was most notable in females (increased by 4.89 times after Eid and by 2.69 times after Ashura), children (increased by 4.69 times after Eid and by 5 times after Ashura) and elderly above the age of 60 years (increased by 5.7 times after Eid and by 3.23 times after Ashura).

**Conclusion:** It is evident that religious events and holidays have important implications on the transmission of SARS-CoV2. This increased in transmission is related mainly to the behavior of the population in those days. Children, female, and elderly were the most affected categories due to these events. A thorough public health plan to limit the spread of the infection at these events should be planned and implemented ahead of time.

## Introduction

Since the outbreak of COVID-19 pandemic different studies have looked on the transmission pattern and risks for transmission of SARS-CoV2. Different risk factors were studied, including biological, demographic, and behavioral factors. SARS-CoV2 is transmitted through respiratory droplets ^(1)^, hence it was evident that behavioral factors were correlated with increased transmission risk, with large in-person gatherings with no social distancing or public health measures carrying the highest risk ^(2)^.

Risk of transmission from an infected case increases with the closeness and duration of contact and is highest with prolonged contact in indoor settings. It has been observed that most local transmissions occur in household contacts, healthcare setting and whenever individuals are in proximity. Risk of transmission also appears high in gatherings ^(3,4,5,6,7)^. Bahrain has recorded the first case in February 24 and the number of new cases have increased since then ^(8)^. The authorities soon placed multiple public health regulations to limit the spread, including banning gatherings, restriction of travel, mandatory face mask wearing, working from home, online schooling and a national testing plan ^(9,10,11,12,13,14)^. The daily recorded cases were noticed to have a correlation with public holidays and social events, especially with Eid and Ashura.

Eid Alfitr is a three-day public holiday in Bahrain. All Muslims celebrate the holiday by having large group prayers in the morning, followed by multiple social gatherings at homes. Like Christmas, it is a festive holiday, and it is common for individuals to visit their families, neighbors, and friends. They would gather for breakfast, lunch, dinner, and short visits to extended family in between. Due to the pandemic, the group prayer was stopped and a public campaign to raise awareness of the importance of staying at home was started.

Ashura is a religious two-day holiday in Bahrain and an event for Muslims. A subset of Muslims has holy religious ceremonies that extend for up to 10 days. Ceremonies are done in large groups and in proximity. However due to the pandemic and banning of indoor gatherings, these ceremonies could continue in small groups outdoors with public health measures.

In this manuscript we study the change in cases and demographics of the newly diagnosed cases before and after two main religious events and holidays in Bahrain: “Eid AlFitr” and “Ashura”.

## Methods

### Study design

This is a national observational study conducted in Bahrain in September 2020 to compare the number and demographics of all newly diagnosed cases before and after Eid Holiday (religious holiday) and Ashura religious event. A 10 days period before the event was compared to a 10 days period after the event by ten days. This period was allowed for the effect of the event to appear on the population. **Figure 1** shows the selected time frames.

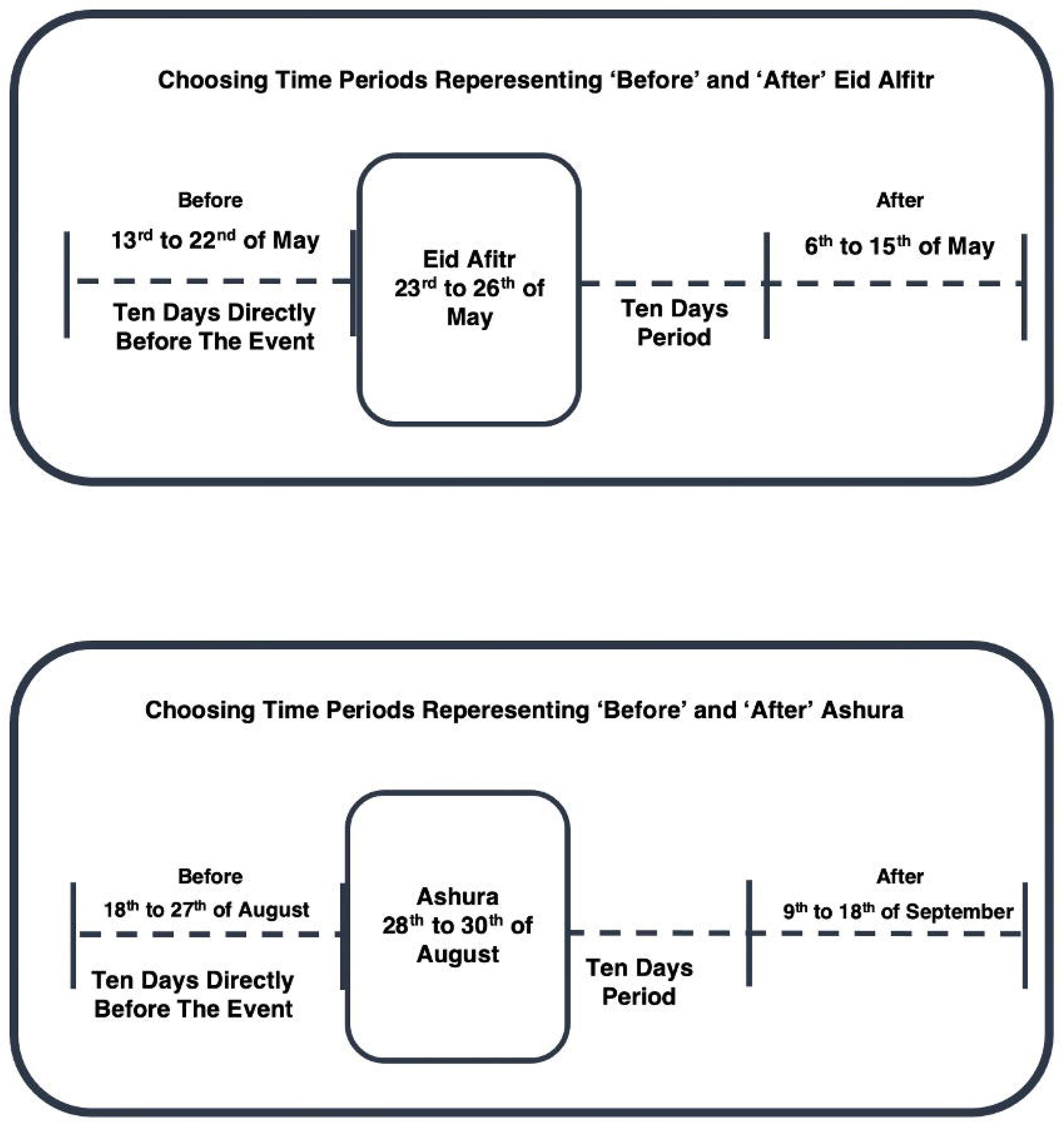

### Testing and diagnosis

A newly diagnosed COVID-19 case was defined as an individual who tested positive for SARS-CoV-2 Envelope (E), RNA-dependent RNA Polymerase (RdRP) and Nucleocapsid (N) genes. The test was conducted by taking a nasopharyngeal (NP) swab. The NP sample was tested for the presence of SARS-CoV-2 by polymerase chain reaction (PCR) analysis. The assay followed the World Health Organization (WHO) approved protocol by Charité Virology ^(15)^ and measured the viral E gene. If the E gene was detected, the sample was subsequently analysed for the SARS-CoV-2 RdRP and N genes. When measuring the initial E gene, a Cycle threshold (Ct) value of >40 was considered negative. Positive and negative controls were included for quality control purposes.

The national testing plan involved testing all suspected cases. All symptomatic individuals and close contacts to a confirmed case were tested. Random community testing was also conducted on high risk areas with potential outbreaks.

### Data collection and Statistical analysis

Data was collected from the National COVID-19 Task Force. Data on the number of tests, number of new cases, case demographics (age, gender, nationality) and presence of symptoms were collected and analyzed. Change in cases was reported as a ratio of the cases after compared to the cases before the event. Ratio above 1 indicate an increase in cases, ratios between 0 and 1 indicated a decrease in cases

Tests of proportion was used to compare proportion of positive tests before and after the events. P-values were considered statistically significant at p<0.05. Data handling, statistical analysis and graphs were done on Microsoft excel (Version 16.4) and STATA (Version 15.1)

### Ethical considerations

Ethical and research approval was obtained from the National COVID19 research and ethics committee (approval code: CRT-COVID2020-081). All methods and analysis of data was approved by the National COVID-19 Research and Ethics Committee and carried out in accordance with the local guideline and ethical guidelines of the Declaration of Helsinki 1975. Informed consent was waived by the Research and Ethical Committee for this study due to the absence of any patient identifying information and the nature of the study design.

## Results

There was significant increase in the number of cases of COVID-19 after both Eid Alfitr and Ashura. Most new cases were found in local Bahrainis. The rise was most notable in females, children, and elderly. There was also a significant increase in symptomatic compared to asymptomatic cases.

### Eid Alfitr

Before Eid Alfitr, the number of tests conducted was 71,553 PCR tests which resulted in 2990 cases with a 4.17% proportion of positive tests. Majority of cases were community acquired infection (98.7%, 2950 cases), while travelers represented a minority (1.3%, 40 cases). Most cases were non-Bahraini (84.2%, 2518 cases) and Males (90.4%, 2703 cases). Asymptomatic cases represented 82.8% of the cases, while symptomatic cases were only 17.2%. Most cases were between the age 20 and 49 years old (84.1%, 2516 cases), while children were 153 cases (5.1% of cases).

After Eid Alfitr, the number of tests conducted was 76,384 PCR tests which resulted in 4987 cases (67% more) with a 6.5% proportion of positive tests. Cases were mainly due to local community transmission (96.9%, 4831 cases). Cases of Bahraini nationals increased by 4.6 times (from 472 cases to 2169 cases) to represent 43.5% of cases. Non-Bahraini cases had a 12% increase in cases. Males continued to represent most cases (71.9% of cases). Female cases increased by 4.8 times (from 287 female cases to 1400 female cases). Symptomatic cases increased by 3.8 times, representing 40% of cases (1995 cases) in that period. The increase in cases was prevalent across all age groups in Bahraini nationals. The most notable increase was in children aged 0 to 9 years old, who increased by 5.5 times from 64 cases to 355 cases(. The elders aged more than 60 years old increased by 5.7 times from 35 cases to 202 cases(. Table 1 summarizes the finding related to Eid AlFitr

**Table (1):** The table demonstrates COVID-19 cases before and after Eid Alfitr. Cases are divided by: transmission source, symptomatic status, gender, nationality, and age. The column on the far right shows the change comparing before and after Eid Alfitr.

|  | 13 to 22 May |  | 6 to 15 June |  | Change |
| --- | --- | --- | --- | --- | --- |
|  | Before Eid Alfitr |  | After Eid Alfitr |  |  |
| Number of tests | 71553 |  | 76384 |  | 1.06x |
| Proportion of positive tests | 4.18% |  | 6.53% |  |  |
| Total cases | 2990 |  | 4987 |  | 1.67x |
| Transmission source |  |  |  |  |  |
| Local cases | 2950 | 98.7% | 4831 | 96.9% | 1.64x |
| Imported cases | 40 | 1.3% | 156 | 3.1% | 3.90x |
| Symptomatic | 514 | 17.2% | 1995 | 40.0% | 3.88x |
| Asymptomatic | 2476 | 82.8% | 2992 | 60.0% | 1.21x |
| Bahraini | 472 | 15.8% | 2169 | 43.5% | 4.60x |
| Non Bahraini | 2518 | 84.2% | 2818 | 56.5% | 1.12x |
| Male | 2703 | 90.4% | 3587 | 71.9% | 1.33x |
| Female | 287 | 9.6% | 1400 | 28.1% | 4.88x |
| Children 0-19 | 153 | 5.1% | 711 | 14.3% | 4.65x |
| Total Non-Bahraini Cases | 2518 |  | 2818 |  | 1.12x |
| Non Bahraini Age Groups |  |  |  |  |  |
| 0-9 | 13 | 0.5% | 72 | 2.6% | 5.54x |
| 10-19 | 14 | 0.6% | 50 | 1.8% | 3.57x |
| 20-29 | 522 | 20.7% | 591 | 21.0% | 1.13x |
| 30-39 | 1130 | 44.9% | 1210 | 42.9% | 1.07x |
| 40-49 | 600 | 23.8% | 602 | 21.4% | 1.00x |
| 50-59 | 191 | 7.6% | 238 | 8.4% | 1.25x |
| 60+ | 48 | 1.9% | 55 | 2.0% | 1.15x |
| Total Bahraini Cases | 472 |  | 2169 |  | 3.60x |
| Bahraini Age Groups |  |  |  |  |  |
| 0-9 | 64 | 13.6% | 355 | 16.4% | 5.55x |
| 10-19 | 62 | 13.1% | 292 | 13.5% | 4.71x |
| 20-29 | 101 | 21.4% | 431 | 19.9% | 4.27x |
| 30-39 | 105 | 22.2% | 405 | 18.7% | 3.86x |
| 40-49 | 58 | 12.3% | 268 | 12.4% | 4.62x |
| 50-59 | 47 | 10.0% | 216 | 10.0% | 4.60x |
| 60+ | 35 | 7.4% | 202 | 9.3% | 5.77x |

The proportion of positive tests was significantly higher after Eid (z = -19.86, p <0.001)

### Ashura

Before Ashura, the number of tests conducted was 97,560 PCR tests which resulted in 3571 cases with a 3.66% proportion of positive tests. The majority of the cases were community acquired infections (99.1%, 3539 cases). The number of cases was 3539 cases, and Bahraini cases constituted 61.6% of the cases (2201 cases). This is a notable rise when compared to Bahraini cases before Eid Alfitr (15.8%) and after Eid Alfitr (43.5%). The male gender represented most cases (60.2%, 2151 cases). Asymptomatic cases represented 73.2%, while symptomatic patients were 26.8%. Cases in Bahrainis were almost evenly spread over all age groups, with children aged 0 to 9 years representing 19.6% of the Bahraini cases.

After Ashura, the number of tests conducted was 118,548 PCR tests which resulted in 7803 cases (2.19 times more) with a 6.58% proportion of positive tests. Cases were almost exclusively due to local transmission (99.3%, 7751 cases). Cases of Bahraini nationals increased by 3.02 times (from 2201 to 6639 cases) to represent 85.1% of cases. Non-Bahraini cases decreased after Ashura (from 1370 to 1164 cases). Female cases increased by 2.69 times, representing 48.9% of all cases almost equivalent to male cases (51.1%). Symptomatic cases increased by 3.37 times, representing 41.3% of cases (3219 cases). The increase in Bahraini cases was close in all age groups, but the major increase was in children aged 0-19 which increased by 5.06 times (from 471 to 2383 cases). Table 2 summarizes the finding related to Ashura.

**Table (2):** The table demonstrates COVID-19 cases before and after Ashura. Cases are divided by: transmission source, symptomatic status, gender, nationality, and age. The column on the far right shows the change comparing before and after Ashura.

|  | 18 to 27 August |  | 9 to 18 September |  | Change |
| --- | --- | --- | --- | --- | --- |
|  | Before Ashura |  | After Ashura |  |  |
| Number of tests | 97560 |  | 118548 |  | 1.22x |
| Proportion of positive tests | 3.66% |  | 6.58% |  |  |
| Total cases | 3571 |  | 7803 |  | 2.19x |
| Transmission Source |  |  |  |  |  |
| Local cases | 3539 | 99.1% | 7751 | 99.3% | 2.19x |
| Imported cases | 32 | 0.9% | 52 | 0.7% | 1.63x |
| Symptomatic | 956 | 26.8% | 3219 | 41.3% | 3.37x |
| Asymptomatic | 2615 | 73.2% | 4582 | 58.7% | 1.75x |
| Bahraini | 2201 | 61.6% | 6639 | 85.1% | 3.02x |
| Non Bahraini | 1370 | 38.4% | 1164 | 14.9% | 0.85x |
| Male | 2151 | 60.2% | 3986 | 51.1% | 1.85x |
| Female | 1420 | 39.8% | 3817 | 48.9% | 2.69x |
| Children 0-19 | 471 | 13.2% | 2383 | 30.5% | 5.06x |
| Total Non-Bahraini Cases | 1370 |  | 1164 |  | 0.85x |
| Non Bahraini Age Groups |  |  |  |  |  |
| 0-9 | 62 | 4.5% | 49 | 4.2% | 0.79x |
| 10-19 | 40 | 2.9% | 23 | 2.0% | 0.58x |
| 20-29 | 297 | 21.7% | 225 | 19.3% | 0.76x |
| 30-39 | 542 | 39.6% | 436 | 37.5% | 0.80x |
| 40-49 | 319 | 23.3% | 261 | 22.4% | 0.82x |
| 50-59 | 87 | 6.4% | 136 | 11.7% | 1.56x |
| 60+ | 23 | 1.7% | 34 | 2.9% | 1.48x |
| Total Bahraini Cases | 2201 |  | 6639 |  | 3.02x |
| Bahraini Age Groups |  |  |  |  |  |
| 0-9 | 431 | 19.6% | 1158 | 17.4% | 2.69x |
| 10-19 | 372 | 16.9% | 1153 | 17.4% | 3.10x |
| 20-29 | 415 | 18.9% | 1163 | 17.5% | 2.80x |
| 30-39 | 353 | 16.0% | 1064 | 16.0% | 3.01x |
| 40-49 | 269 | 12.2% | 855 | 12.9% | 3.18x |
| 50-59 | 197 | 9.0% | 717 | 10.8% | 3.64x |
| 60+ | 164 | 7.5% | 529 | 8.0% | 3.23x |

The proportion of positive tests was significantly higher after Ashura (z = -30.26, p <0.001)

## Discussion

After COVID-19 emerged in Bahrain, the government placed multiple public health regulations to contain the outbreak ^(14)^. Due to these precautionary measures, many Bahrainis stayed at home before Eid Alfitr ^(16)^. This could explain the low number of Bahraini cases (472 cases) back then.

Because of the cultural nature of Eid Alfitr and Ashura, it is no surprise to see the increase in incidence after each event. In both events, the increase in cases was more prominent in Bahrainis because they partake in these events much more than non-Bahrainis. Eid have resulted in 1697 new cases within Bahraini nationals (3.6 times more) and Ashura have resulted in 4438 cases more (3.02 times more)

Both events resulted in an increase in the number of cases across all age groups. The largest margin of increase was witnessed in the extremes of age: 0-19 years old and elderly above 60 years old. With the implementation of “staying at home” and closure of schools, mosques and majlis (indoor gatherings with majority of attendees being elders) children aged 0 to 19 and elders 60 years and older spent most, if not all, of their time at home. This greatly limited the number of cases in these age groups before Eid Alfitr: (153) children and (82) elderlies. At that time, most Bahraini cases were aged 20 to 49. These were working people who still had to attend work and encounter different people, leading them to have a higher risk of COVID-19.

When Eid Alfitr came, it was observed that many families and friends gathered indoors, despite advice to quarantine. Furthermore, it is common for smaller families to gather with similar sized families and visit each other in one big gathering that is usually centered around the elderly of the family. This led to a prominent increase in Bahraini cases. The greatest rise observed was in children, females’ elders aged 60 and above. This is likely due to the abstinence of children, females, and the elderly from much social interaction before Eid Alfitr.

Both Eid Alfitr and Ashura increased the number of cases for both genders greatly. Females exhibited a more drastic increase in percentage of total cases when compared in the before and after periods of both events. Which may suggest that the increase of infection rate cannot attributed to a single gender, since both participated in the events. The general higher number in males could be explained by the higher male to female ration in Bahrain ^(17)^. This could also be explained by gender differences in perceived risk- and risk-taking behaviors described by Byrnes et al ^(18)^.

At all times observed, most non-Bahraini cases were aged between 20 and 49. Most of the non-Bahrainis infected were migrant labor workers, typically in the age group mentioned ^(19)^. The large number of cases in the labor workers could be attributed to poor health education and crowded living conditions. Bahraini cases were the minority of the cases during the beginning of the pandemic. Eid Alfitr could be described as the turning point, where cases distribution in Bahrain tipped towards the Bahraini population. Due to the closeness and nature of the Bahraini culture this tip in the scales escalated and created the increase between Bahraini and Non-Bahrainis cases observed. The increase in proportion of Bahraini to non-Bahraini cases remained consistent until the present day.

It is important to note that both after Eid Alfitr and after Ashura the number of tests has increased by (7%) and (22%), respectively. The number of total cases also increased at both time periods at a much larger extent. Eid holidays have resulted in 1697 cases more when compared to the period before that (1.67 times more). Ashura have resulted in 7803 cases in a 10 days period (2.19 times more). The small increase in testing accompanied by the high increase in total cases indicate that Eid Alfitr and Ashura directly affected the incidence and resulted in higher transmission, and that these results could not have been simply due to more testing alone. This was more apparent when the proportion of positive tests was compared. The increase in number of tests after these events was due to an increase in testing demand. As it was evident that the number of symptomatic cases increased, which also led to an increase in contact tracing and testing too.

The increase in cases and the proportion of positive tests observed in the period after Eid Alfitr was brought down with the measures enforced by a campaign aimed to increase the general awareness and change the behavior in the Bahraini population as a whole. The campaign included posting comparison of cases before and after Eid and posting anonymized contract tracing result which showed the extent of infections in household members ^(20)^. This decrease was clear, as it can be observed by the number of cases before the events of Ashura (Table 2). As the period of Ashura started, it contributed greatly to increasing the number of cases and increasing outbreaks in different parts of the country

Before Ashura, the total cases diagnosed were 3539. This was an increase compared to cases before Eid Alfitr (2990 cases). This is a new baseline in cases which can be explained by the change in people’s behavior, especially following reopening measures. However, cases after Eid Alfitr were still higher (4831 cases), implying the significant effect Eid Alfitr had on increasing transmission.

In Ashura, the participation in ceremonies were separated by gender. However, this separation resulted in many small indoor gatherings in proximity with a preacher reciting speeches and chants. These ceremonies extend from two to ten hours per day for two to ten days Tang et al ^(21)^ stated that indoor gatherings with a person performing or in this case reciting speeches and chants may enable the transmission of the virus through aerosolization and not only droplets. Making such events as superspreading events. This was also reported in a choir rehearsal where 53 out 61 attendees were infected despite adequate precaution for fomite and droplet transmission taken and none of the attendees exhibited symptoms ^(22,23)^. Miller et al also stated that “this indoor transmission risk may have been increased because of high occupancy, long duration, loud vocalization, and poor ventilation” ^(22)^.

Eid Alfitr effect on the number of cases was largely due to the festive nature of the holiday, where many individuals visit their families, extended families, and friends. Eid Alfitr is celebrated by all Muslim. In comparison, only a subset of Muslims holds religious ceremonies and gatherings in Ashura. The fact that Ashura resulted in such increase in the number of cases among all age groups, especially children and females, can be attributed to the nature of the ceremonies conducted during Ashura.

After both events, symptomatic cases have increased. This could be explained by the higher incidence of COVID-19 in older individual, as it has been reported that younger patients were more likely to be asymptomatic ^(24)^. The more severe and symptomatic cases could also be due to different viral strains that have emerged from different clusters due to gatherings, and this might be worth exploring with genomic sequencing.

Public health measures and guidance such as: banning indoor gatherings, online ceremonies and limiting capacity in the participation in the ceremonies were put in place to limit the effect of the religious event. As new cases emerged and contact tracing was done, it was found that people did not adhere to recommended public health safety measures. Many gatherings in proximity were conducted which increased the risk for transmission between the population. Moreover, during these gatherings meals were exchanged between gatherings and individuals which have contributed to the increase in infection rate.

Furthermore, it was observed that due to the decision of limiting the Ashura ceremonies and conducting them online, this has caused many of the small gatherings. As people could not participate in person, many decided to gather in houses of relatives and friends to attend the ceremonies and share the spirituality of Ashura. Which may suggest that providing an online option for something that required a large social gathering may result in many smaller gatherings. One of the large contributors to the spread of infection in these gatherings were individuals that visited multiple gatherings. Such individuals would visit one small gathering for a few hours and then leave to attend a different gathering. This has resulted in gatherings that are at extremely high risks for transmission. Another factor that needs to be considered is the duration of events. Each gathering would last for a minimum of 2 hours and could last up to 6-10 hours on daily bases for the whole period of Ashura. Thus, creating an environment where one positive case could infect more than it can under normal circumstances.

Such events are of extreme importance to people, whether they are ceremonies or family gatherings. Implementing rules and regulation to limit these gatherings is required, however not sufficient. A pre-planned public health plan should be started in advance of these events to raise the public awareness on the risk of gatherings and to improve the public compliance. Moreover, it is also equally important to raise awareness on how to decrease transmission in such events or gatherings, as it is evident that a large number of the population will continue to gather. Personal hygiene, physical distancing, wearing face masks, time limitation and proper ventilation are all measures that can help reduce the risk of transmission in such events and gatherings. ^(22, 25)^. This could be a crucial measure to decrease the transmission and help fight the pandemic.

### Conclusion

It is evident that religious events and holidays have important implications on the transmission of SARS-CoV2. This increased in transmission is related mainly to the behavior of the population in those days. Children, female, and elderly were the most affected categories due to these events. Symptomatic individuals increased after these events. These events are considered high risk for transmission and should be included in models forecasting the number of cases. A thorough public health plan to limit the spread of the infection at these events should be planned and implemented ahead of time. The plan should include raising public awareness prior to the event, limiting the events, proposing a safer substitute, involving religious leaders, and enhancing measures to decrease the risk of transmission within an event.

## Data Availability

Available on reasonable request

## Declaration

### Ethics approval

Approval granted after review from National COVID-19 Research and Ethics Committee

### Consent to participate

Not applicable

### Consent for publication

All author approved to publish this data Availability of data and material: Available on reasonable request Code availability: All data were entered in Microsoft excel

### Authors’ contributions

AIA, AA, MAM involved in data collection, analysis of the data and initial manuscript preparation. MAQ reviewed the manuscript. MAQ, AA developed the idea and final drafting of the manuscript.

## References

1. COVID-19 Overview and Infection Prevention and Control Priorities in non-US Healthcare Settings: Centers for Disease Control and Prevention; 2020 [Available from: https://www.cdc.gov/coronavirus/2019-ncov/hcp/non-us-settings/overview/index.html.

2. People With Certain Medical Conditions: Centers for Disease Control and Prevention; 2020 [Available from: https://www.cdc.gov/coronavirus/2019-ncov/need-extra-precautions/people-with-medical-conditions.html.

3. Wang D, Hu B, Hu C, Zhu F, Liu X, Zhang J, et al. Clinical Characteristics of 138 Hospitalized Patients With 2019 Novel Coronavirus-Infected Pneumonia in Wuhan, China.JAMA.2020;323(11):1061–9.

4. Luo L, Liu D, Liao X, Wu X, Jing Q, Zheng J, et al. Contact Settings and Risk for Transmission in 3410 Close Contacts of Patients With COVID-19 in Guangzhou, China : A Prospective Cohort Study. Ann Intern Med. 2020.

5. Bi Q, Wu Y, Mei S, Ye C, Zou X, Zhang Z, et al. Epidemiology and transmission of COVID-19 in 391 cases and 1286 of their close contacts in Shenzhen, China: a retrospective cohort study. Lancet Infect Dis.2020;20(8):911–9.

6. COVID-19 National Emergency Response Center EaCMT, K.rea Centers for Disease Control and Prevention. Coronavirus Disease-19: Summary of 2,370 Contact Investigations of the First 30 Cases in the Republic of Korea. Osong Public Health Res Perspect.2020;11(2):81–4.

7. Karami M, Doosti-Irani A, Ardalan A, Gohari-Ensaf F, Berangi Z, Massad E, et al. Public Health Threats in Mass Gatherings: A Systemic Review. Disaster Medicine and Public Health Preparedness.2019;13(5-6):1035–46.

8. Ministry of Health: Patient with Coronavirus (COVID-19) isolated and receiving treatment: Ministry of Health; 2020 [Available from: https://www.moh.gov.bh/COVID19/Details/3753.

9. All public and private schools and kindergartens to be closed for two weeks Ministry of Health; 2020 [Available from: https://www.moh.gov.bh/COVID19/Details/3879.

10. All Universities and Higher Education institutions closed for two weeks: Ministry of Health 2020 [Available from: https://www.moh.gov.bh/COVID19/Details/3874.

11. Civil Aviation Affairs suspends all flights from Dubai, Sharjah for 48 hours: Ministry of Health 2020 [Available from: https://www.moh.gov.bh/COVID19/Details/3820.

12. The Ministry of Health to arrange medical examinations for all who have visited Iran during February: Ministry of Health 2020 [Available from: https://www.moh.gov.bh/COVID19/Details/3788.

13. Foreign Ministry calls upon all citizens not to travel during this period unless absolutely necessary: Ministry of Health; 2020 [Available from: https://www.moh.gov.bh/COVID19/Details/3924.

14. Coronavirus TNMTFtCt. The Decisions of the Government Executive Committee chaired by HRH the Crown Prince, Deputy Supreme Commander and First Deputy Prime Minister 2020 [Available from: https://www.moh.gov.bh/Content/Upload/File/637202407881866000-DecisionsGECEn.pdf.

15. Corman V, Bleicker T, Brünink S, Drosten C, Landt O, Koopmans M, et al. Diagnostic detection of 2019-nCoV by real-time RT-PCR2020. Available from: https://www.who.int/docs/default-source/coronaviruse/protocol-v2-1.pdf.

16. COVID-19 Mobility Report2020. Available from: https://www.gstatic.com/covid19/mobility/2020-06-07_BH_Mobility_Report_en.pdf.

17. Bahrain in Figures: Information and eGovernment Authority; 2017. Available from: https://www.iga.gov.bh/Media/Pdf-Section/Bahrain_in_figures_Booklet.pdf.

18. Byrnes JP, Miller DC, Schafer WD. Gender differences in risk taking: A meta-analysis. Psychological Bulletin.1999;Vol 125(3):367–83.

19. Bahrain Labour Market Indicators2019; (Issue 46). Available from: http://lmra.bh/portal/files/cms/shared/file/Newsletter/Newsletter(Q2_2019)-English.pdf.

20. Contact Tracing2020. Available from: https://www.moh.gov.bh/COVID19/ContactTracing1.

21. Tang S, Mao Y, Jones RM, Tan Q, Ji JS, Li N, et al. Aerosol transmission of SARS-CoV-2? Evidence, prevention and control. Environ Int.2020;144:106039.

22. Miller SL, Nazaroff WW, Jimenezz JL, Boerstra A, Buonanno G, Dancer SJ, et al. Transmission of SARS-CoV-2 by inhalation of respiratory aerosol in the Skagit Valley Chorale superspreading event 2020. Available from: https://www.medrxiv.org/content/10.1101/2020.06.15.20132027v2.

23. Read R. Coronavirus Choir Outbreak2020. Available from: https://www.latimes.com/world-nation/story/2020-03-29/coronavirus-choir-outbreak.

24. Li Y, Shi J, Xia J, Duan J, Chen L, Yu X, et al. Asymptomatic and Symptomatic Patients With Non-severe Coronavirus Disease (COVID-19) Have Similar Clinical Features and Virological Courses: A Retrospective Single Center Study. Front Microbiol.2020;11:1570.

25. Sugano N, Ando W, Fukushima W. Cluster of SARS-CoV-2 infections linked to music clubs in Osaka, Japan: asymptomatically infected persons can transmit the virus as soon as 2 days after infection. J Infect Dis. 2020.

